# Adaptive Deep Brain Stimulation in Parkinson’s Disease: A Delphi Consensus Study

**DOI:** 10.1101/2024.08.26.24312580

**Authors:** M. Guidetti, T. Bocci, M. De Pedro Del Álamo, G. Deuschl, A. Fasano, R. Martinez Fernandez, C. Gasca-Salas, C. Hamani, J.K. Krauss, A. A. Kühn, P. Limousin, S. Little, A.M. Lozano, N.V. Maiorana, S. Marceglia, M.S. Okun, S. Oliveri, J. L. Ostrem, E. Scelzo, A. Schnitzler, P.A. Starr, Y. Temel, L. Timmermann, G. Tinkhauser, V. Visser-Vandewalle, J. Volkmann, A. Priori

## Abstract

**Importance:** If history teaches, as cardiac pacing moved from fixed-rate to on-demand delivery in in 80s of the last century, there are high probabilities that closed-loop and adaptive approaches will become, in the next decade, the natural evolution of conventional Deep Brain Stimulation (cDBS). However, while devices for aDBS are already available for clinical use, few data on their clinical application and technological limitations are available so far. In such scenario, gathering the opinion and expertise of leading investigators worldwide would boost and guide practice and research, thus grounding the clinical development of aDBS.

**Observations:** We identified clinical and academically experienced DBS clinicians (n=21) to discuss the challenges related to aDBS. A 5-point Likert scale questionnaire along with a Delphi method was employed. 42 questions were submitted to the panel, half of them being related to technical aspects while the other half to clinical aspects of aDBS. Experts agreed that aDBS will become clinical practice in 10 years. In the present scenario, although the panel agreed that aDBS applications require skilled clinicians and that algorithms need to be further optimized to manage complex PD symptoms, consensus was reached on aDBS safety and its ability to provide a faster and more stable treatment response than cDBS, also for tremor-dominant Parkinson’s disease patients and for those with motor fluctuations and dyskinesias.

**Conclusions and Relevance:** Despite the need of further research, the panel concluded that aDBS is safe, promises to be maximally effective in PD patients with motor fluctuation and dyskinesias and therefore will enter into the clinical practice in the next years, with further research focused on algorithms and markers for complex symptoms.

## 1. INTRODUCTION

Deep Brain Stimulation (DBS) is a standard neurosurgical therapy to treat selected patients with neurological disorders including essential tremor (ET), Parkinson’s disease (PD), and dystonia.^1^ Traditionally, DBS has been employed using open-loop stimulation techniques, i.e., delivering continuous, uninterrupted stimulation at the same parameter setting (conventional DBS, cDBS) that is independent of the real-time patient’s functional status or of the side effects induced by intermittent stimulation. In PD, DBS of the subthalamic nucleus (STN-DBS^2^), has been prominently associated with stimulation-induced speech impairments,^3^ risk of falling,^4^ dyskinesia,^5^ stimulation-induced impulsivity,^6^ and, more importantly, only partial control of clinical fluctuations.^7^ Adaptive DBS (aDBS) was conceived to overcome some of the disadvantages of cDBS by facilitating optimized current delivery to improve symptoms and drive improved outcomes.^8^ This technology relies on the principle of on-demand or contingency-based stimulation, where clinically relevant biofeedback signals (e.g., brain signals) can be used to determine more effective characteristics of the stimulation (or changes to other parameters) to be delivered in real-time in order to address emerging symptoms or side effects.^9^ Currently, in the field of movement disorders,^8^ both electrocorticographic signals registered from cortical electrode strips and local field potentials (LFPs) recorded directly from the DBS electrodes have been explored in feasibility testing.^8,10^

Although the aDBS concept is perceived as a natural evolution of current cDBS, in line with the historical development of cardiac pacemakers, the evidence collected on its clinical application needs to be expanded, especially to better understand the emerging limitations, and to boost its adoption and understanding in everyday clinical practice. For instance, in PD, where beta band STN LFPs can be applied as control signal for DBS amplitude adjustments,^11^ experiments revealed an inconsistent correlation to clinical outcome scores on validated scales of PD disability and motor dysfunction,^12,13^ especially with patients presenting with different phenotypes (e.g., tremor dominant or akinetic rigid PD).^14^ Therefore, some experts suggested that LFP power alone might not provide a reliable biomarker for aDBS^15^ because of the failure to represent the complex pathological cortical-subcortical circuital activity in PD and, in turn, to serve as a robust marker, particularly for complex symptoms.^16–18^

Such a challenging scenario demands for the integration of the knowledge derived from clinical data and from the experience of leading experts that will serve to (1) provide a clear scenario for aDBS advantages and limitations at the current state-of-the-art, (2) guide the future design of trials and (3) highlight the most promising directions for aDBS. To boost this dialogue, we identified internationally recognized clinical and academic DBS experts to discuss the methodological and clinical challenges and we asked them to participate in a Delphi method-based study.^19^

## 2. METHODS

The Delphi study methodology is a multistage process designed to combine opinions into group consensus,^20^ where a series of structured questionnaires (rounds) are anonymously completed by experts (panellists) and the responses from each questionnaire fed back in summarised form to the participants.^21–23^ This allows the panellists to reassess their initial judgments, considering the positive aspects of interacting groups (e.g., inclusion of different backgrounds) without the negative ones (e.g., influence of dominant members).^24^ For the purpose of our study, a modified Delphi process^25,26^ was designed in three rounds, which are considered as sufficient to collect the needed information and to reach a consensus.^21,24,27,28^ A Steering Committee (SC) of experts (n=8) based on the collaborative network of the leading authors discussed the topic and developed a structured questionnaire including key items pertinent to aDBS using five-point Likert scales (1=strongly disagree; 2=disagree; 3=undecided; 4=agree; 5=strongly agree).^19^ In rounds one, two and three, quantitative assessments to reach the consensus were performed by SC members and a larger Expert Panel (EP, n=13). Therefore, a total of 21 panellists took part in the assessment, which is a number of experts within the recommended range.^24,29^ Since no exact criterion is currently available on the definition of “expert”,^30^ we decided to consider positional leaders in the field, as suggested by previous works.^31^ The panellists were asked to rate 42 statements on several technical (21 statements) and clinical (21 statements) aspects of aDBS (Table 1). In order to maintain the rigor of this method, we considered a response rate of >70% for each round^32^ to be a minimum. Electronic questionnaires were used in all steps of the process. In case one item reached a consensus during the first or second round, it was excluded from the following round to avoid confirmation bias.

**Table 1.** Five-point Likert questionnaire with the results (median ± IQR) for each round.

| Statement | 1<sup>st</sup> round<br>(n=19;<br>RR=90.5%) | 2<sup>nd</sup> round<br>(n=20;<br>RR=95.2%) | 3<sup>rd</sup> round<br>(n=20;<br>RR=95.2%) |
| --- | --- | --- | --- |
| **Technical aspects of adaptive DBS** |  |  |  |
| **S1.** Adaptive DBS is at the beginning of its clinical applications, but I think that there may still be technological limitations | 4 ± 1 | 4 ± 0.25 | **4 ± 0 – C.R.** |
| **S2.** I think that a possible limitation of the diffusion of adaptive DBS are high costs | 3 ± 1 | 3 ± 1.25 | 3 ± 1 |
| **S3.** I think adaptive DBS is applicable in patients with not well-positioned electrodes | 1 ± 1 | 1 ± 1 | **1 ± 0 – C.R.** |
| **S4.** I think adaptive DBS is applicable when one side only is able to record | 3 ± 1 | 4 ± 1 | 4 ± 1 |
| **S5.** I think that only modulating the amplitude might be a limiting factor of adaptive DBS | 3 ± 2 | 2 ± 2 | 2.5 ± 2 |
| **S6.** I think an actual risk for adaptive DBS is overstimulation | 3 ± 1 | 3 ± 1 | 3 ± 0 |
| **S7.** I think an actual risk for adaptive DBS is under stimulation | 3 ± 1.5 | 3 ± 1 | 3 ± 1 |
| **S8.** I think adaptive DBS requires high level of expertise | 4 ± 1 | 5 ± 1 | **5 ± 0 – C.R.** |
| **S9.** I think adaptive DBS is feasible only in experienced DBS centres with neurophysiological expertise | 4 ± 1.5 | 4 ± 0.25 | 4 ± 0 |
| **S10.** I think adaptive DBS surgery is time-consuming | 3 ± 2 | 4 ± 2 | 4 ± 2 |
| **S11.** I think adaptive DBS programming is time-consuming | 4 ± 3 | 4 ± 1 | 4 ± 1 |
| **S12.** I think that automatic programming will reduce programming time | 5 ± 1 | 5 ± 1 | 5 ± 1 |
| **S13.** I think that automatic programming is safe as long as the neurologist can set upper and lower limits for stimulation intensity | 4 ± 0 | **4 ± 0 – C.R.** | - |
| **S14.** I think fast adaptation adaptive DBS methods are superior to slow adaptation adaptive DBS methods | $3 \pm 1$ | $3 \pm 0$ | **$3 \pm 0 - \text{C.R.}$** |
| **S15.** I think slow adaptation adaptive DBS methods are superior to fast adaptation adaptive DBS methods | $3 \pm 1$ | $3 \pm 0$ | **$3 \pm 0 - \text{C.R.}$** |
| **S16.** I think adaptive DBS will be based more likely on feedback from wearables than on signal recording from the DBS electrodes | $2 \pm 1$ | $2 \pm 0$ | $2 \pm 0.25$ |
| **S17.** I think adaptive DBS will be based more likely on signal recording from the DBS electrodes than on feedback from wearables | $4 \pm 1$ | $4 \pm 1$ | $4 \pm 0$ |
| **S18.** I think adaptive DBS would help to diffuse DBS with segmented electrodes | $3 \pm 1$ | $3 \pm 0$ | **$3 \pm 0 - \text{C.R.}$** |
| **S19.** I think the rapid development of artificial intelligence (AI) will fuel the clinical use of adaptive DBS | $4 \pm 1$ | $4 \pm 1$ | $4 \pm 0.25$ |
| **S20.** I think current pacemaker technology in principle allows to install adaptive DBS algorithms | $4 \pm 0.5$ | $4 \pm 0.25$ | **$4 \pm 0 - \text{C.R.}$** |
| **S21.** I think changes in technology are still necessary to foster adaptive DBS soon | $4 \pm 1$ | $4 \pm 1$ | $5 \pm 1$ |
| **Clinical aspects of Adaptive DBS** |  |  |  |
| **S22.** I think adaptive DBS will be clinical routine in 10 years from now | $4 \pm 0$ | $4 \pm 1$ | **$4 \pm 0 - \text{C.R.}$** |
| **S23.** I think adaptive DBS will be clinical routine in 5 years from now | $3 \pm 1.5$ | $3 \pm 1$ | $3 \pm 1$ |
| **S24.** The side effects (ramping) will lead to many patients being unable to tolerate adaptive DBS | $2 \pm 1$ | $2.5 \pm 1$ | $2.5 \pm 1$ |
| **S25.** I think adaptive DBS is a safe technology | $4 \pm 0.5$ | $4 \pm 0$ | **$4 \pm 0 - \text{C.R.}$** |
| **S26.** I think adaptive DBS is applicable on a large scale | $3 \pm 1$ | $3 \pm 1$ | $3 \pm 1$ |
| **S27.** I think adaptive DBS is applicable only for non-tremor patients with Parkinson's | $2 \pm 1$ | $2 \pm 0.25$ | $2 \pm 1$ |
| disease |  |  |  |
| **S28.** I think adaptive DBS is applicable also for tremor-dominant patients with Parkinson's disease | $4 \pm 0.5$ | **$4 \pm 0 - \text{C.R.}$** | - |
| **S29.** I think the primary clinical indication for adaptive DBS will rather be tremor than Parkinson's disease | $2 \pm 1$ | $2 \pm 1$ | $2 \pm 0$ |
| **S30.** I think the patient profile who will likely benefit from adaptive DBS is the patient with significant motor fluctuations before DBS | $4 \pm 1.5$ | $4 \pm 1.25$ | **$4 \pm 0 - \text{C.R.}$** |
| **S31.** I think the patient profile who will likely benefit from adaptive DBS is the patient with significant motor fluctuations on conventional DBS | $4 \pm 0$ | $4 \pm 0$ | **$4 \pm 0 - \text{C.R.}$** |
| **S32.** I think the patient profile who will likely benefit from adaptive DBS is the patient with significant dyskinesias on conventional DBS | $4 \pm 1.5$ | $4 \pm 1$ | **$4 \pm 0 - \text{C.R.}$** |
| **S33.** I think that adaptive DBS will improve non-motor aspects of Parkinson's disease | $3 \pm 1$ | $3 \pm 1$ | $3.5 \pm 1$ |
| **S34.** I think that adaptive DBS will reduce stimulation induced side effects | $4 \pm 1$ | $4 \pm 0.25$ | $4 \pm 0$ |
| **S35.** I think the long-term impact of adaptive DBS might be positive for the patients | $4 \pm 0.5$ | $4 \pm 1$ | **$4 \pm 0 - \text{C.R.}$** |
| **S36.** I think adaptive DBS might more easily adapt to pharmacological changes | $4 \pm 1$ | $4 \pm 1$ | $4 \pm 0$ |
| **S37.** I think adaptive DBS leads to faster stable treatment response after DBS surgery once a setting is defined | $4 \pm 1$ | $4 \pm 1$ | **$4 \pm 0 - \text{C.R.}$** |
| **S38.** I think fast adaptation adaptive DBS leads to long term plastic changes | $3 \pm 1$ | $3 \pm 0.25$ | **$3 \pm 0 - \text{C.R.}$** |
| **S39.** I think adaptive DBS will improve patient's well-being because adaptive DBS automatically increases stimulation if patient forgets to take medication | $3 \pm 1.5$ | $4 \pm 1$ | $4 \pm 1$ |
| **S40.** I think adaptive DBS will improve patient's well-being because adaptive DBS automatically decreases stimulation if patient accidentally takes too high a dose of | $4 \pm 1$ | $4 \pm 1$ | $4 \pm 1$ |
| medication |  |  |  |
| **S41.** I think adaptive DBS decreases the number of patient visits to neurologists for programming | $3 \pm 1.5$ | $3 \pm 2$ | $3 \pm 0.25$ |
| **S42.** I think adaptive DBS makes medication titration easier – with less precision required | $3 \pm 1$ | $3 \pm 0.25$ | $3 \pm 0.25$ |

Delphi Panel members were asked to rate their agreement with each statement (1=strongly disagree; 2=disagree; 3=undecided; 4=agree; 5=strongly agree). R.R. = response rate; C.R. = consensus reached; PD = Parkinson's disease; DBS = deep brain stimulation.

Although no guidelines are available,^30^ consensus was achieved when ≥80% of the responses fell in the same response label.^19,33^ Data were analysed and reported by descriptive statistics. We opted for median and interquartile range (IQR), as suggested by the literature.^24,34–36^ We report the results of each round separately in both textual (i.e., with median ± IQR) and graphical representation, to better illustrate the strength of support for each round.^30^

## 3. RESULTS

### 3.1. Specialists panel

For the SC, all the eight invited authors agreed to participate (SC=8, response rate: 100%). For the EP, out of the 20 authors identified, two declined to participate and five did not reply (EP=13, response rate: 65%). Therefore, the overall number of panellists was 21 (overall response rate: 75%, see eTable 1 in Supplementary Materials). Demographic characteristics of the panellists are displayed in Table 2. Briefly, most of them were male (16, 76%), >50 years old (14, 66.6%) and high-experienced in clinical routine (20, 95.5% with >10 years of clinical experience) and research (19, 90.4% and 18, 85.7% with >10 years of experience in, respectively, the DBS field and DBS clinical trials) settings.

**Table 2.**
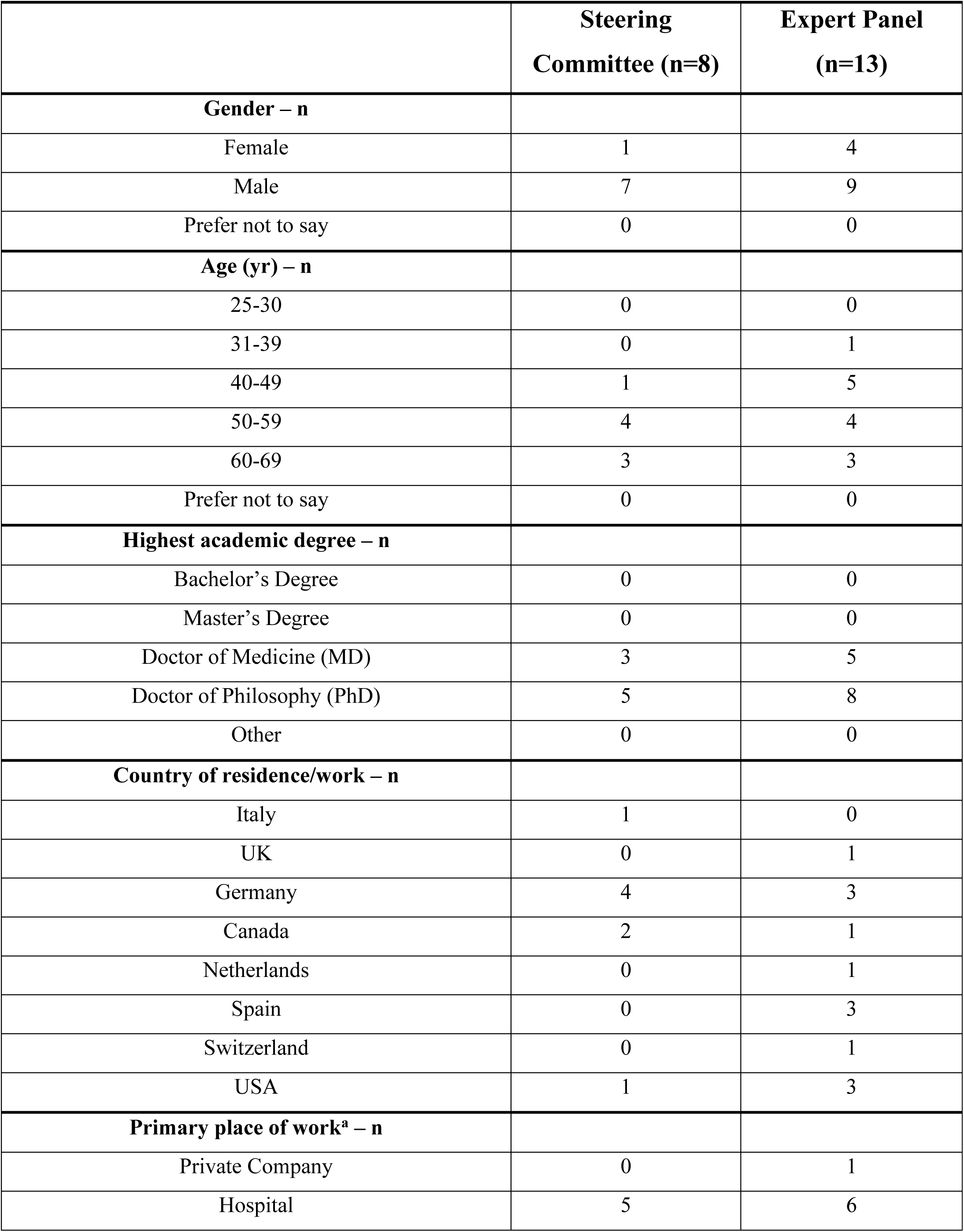
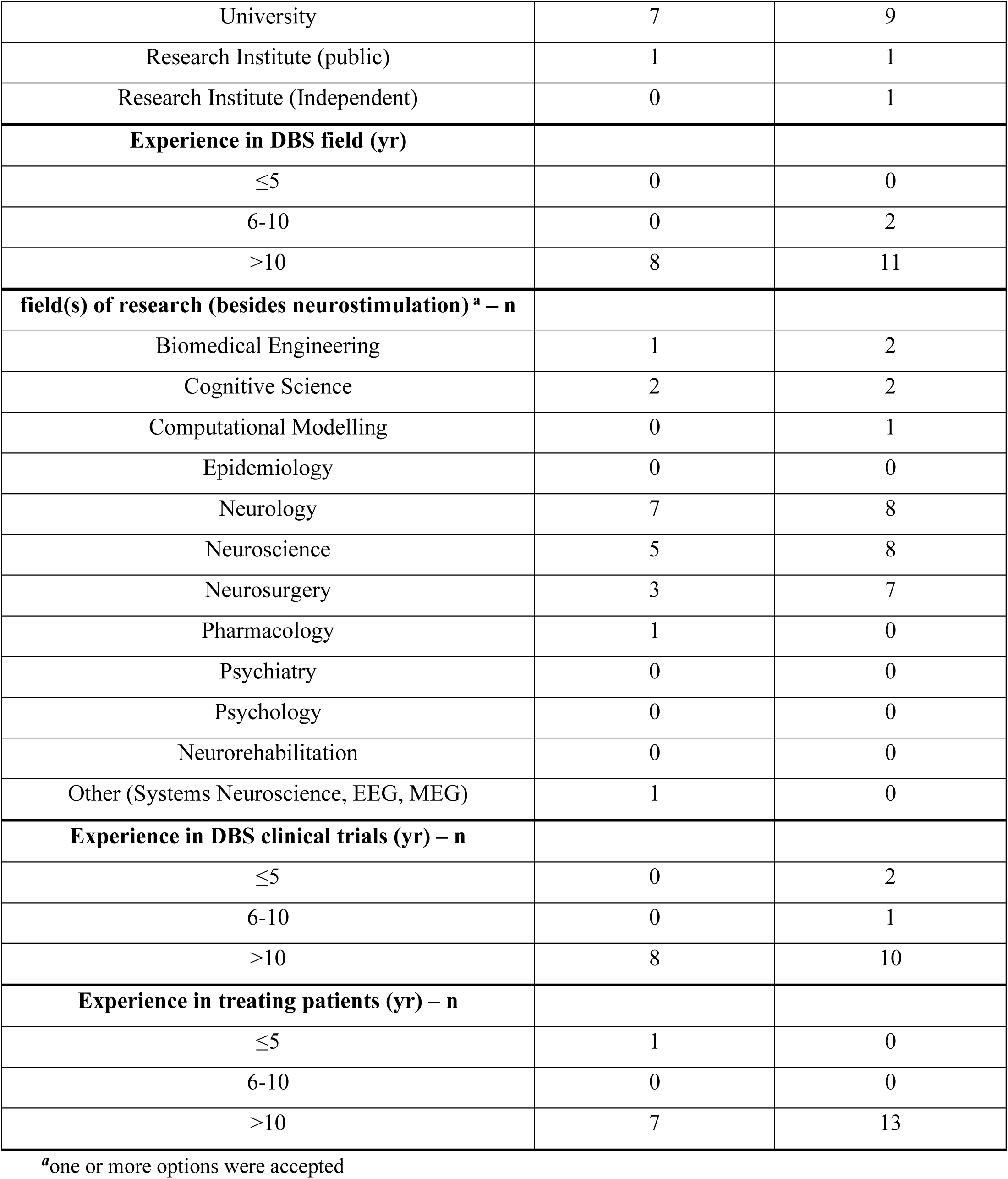
Demographic and academic information for the Delphi Panel members.

|  | **Steering**<br>**Committee (n=8)** | **Expert Panel**<br>**(n=13)** |
| --- | --- | --- |
| **Gender – n** |  |  |
| Female | 1 | 4 |
| Male | 7 | 9 |
| Prefer not to say | 0 | 0 |
| **Age (yr) – n** |  |  |
| 25-30 | 0 | 0 |
| 31-39 | 0 | 1 |
| 40-49 | 1 | 5 |
| 50-59 | 4 | 4 |
| 60-69 | 3 | 3 |
| Prefer not to say | 0 | 0 |
| **Highest academic degree – n** |  |  |
| Bachelor's Degree | 0 | 0 |
| Master's Degree | 0 | 0 |
| Doctor of Medicine (MD) | 3 | 5 |
| Doctor of Philosophy (PhD) | 5 | 8 |
| Other | 0 | 0 |
| **Country of residence/work – n** |  |  |
| Italy | 1 | 0 |
| UK | 0 | 1 |
| Germany | 4 | 3 |
| Canada | 2 | 1 |
| Netherlands | 0 | 1 |
| Spain | 0 | 3 |
| Switzerland | 0 | 1 |
| USA | 1 | 3 |
| **Primary place of work<sup>a</sup> – n** |  |  |
| Private Company | 0 | 1 |
| Hospital | 5 | 6 |
| University | 7 | 9 |
| Research Institute (public) | 1 | 1 |
| Research Institute (Independent) | 0 | 1 |
| **Experience in DBS field (yr)** |  |  |
| ≤5 | 0 | 0 |
| 6-10 | 0 | 2 |
| >10 | 8 | 11 |
| **field(s) of research (besides neurostimulation)<sup>a</sup> – n** |  |  |
| Biomedical Engineering | 1 | 2 |
| Cognitive Science | 2 | 2 |
| Computational Modelling | 0 | 1 |
| Epidemiology | 0 | 0 |
| Neurology | 7 | 8 |
| Neuroscience | 5 | 8 |
| Neurosurgery | 3 | 7 |
| Pharmacology | 1 | 0 |
| Psychiatry | 0 | 0 |
| Psychology | 0 | 0 |
| Neurorehabilitation | 0 | 0 |
| Other (Systems Neuroscience, EEG, MEG) | 1 | 0 |
| **Experience in DBS clinical trials (yr) – n** |  |  |
| ≤5 | 0 | 2 |
| 6-10 | 0 | 1 |
| >10 | 8 | 10 |
| **Experience in treating patients (yr) – n** |  |  |
| ≤5 | 1 | 0 |
| 6-10 | 0 | 0 |
| >10 | 7 | 13 |

<sup>a</sup>one or more options were accepted

### 3.2. Delphi Panel results

As for the 21 statements on the technical aspects of aDBS, the first round led to no consensus for any of the statements (see eFigure 1 in Supplementary Materials); in the second, the consensus was reached in only one statement (see eFigure 2 in Supplementary Materials); finally, in the third round, consensus was reached in other seven statements, for a total of eight out of 21 statements (see fig.1). More specifically, in the second round, the panellists agreed that automatic programming would be safe as long as stimulation intensity is constrained by upper and lower limits (90% agreed, median ± IQR: 4 ± 0). After the third round, panellists agreed that aDBS has technological limitations (Statement 1 – 80% agreed, median ± IQR: 4 ± 0), but that current pacemaker technology might be suitable to implement aDBS algorithms (Statement 20 – 90% agreed, median ± IQR: 4 ± 0). They strongly agreed that it requires high levels of expertise (statement 8 – 80% strongly agreed, median ± IQR: 5 ± 0), but strongly disagreed in its feasibility for patients with not well-positioned electrodes (statement 3 – 85% strongly disagreed, median ± IQR: 1 ± 0). Lastly, panellists were undecided on the role of aDBS in spreading segmented electrodes use (Statement 18 – 85% undecided, median ± IQR: 3 ± 0), or whether fast adaptation methods are superior or inferior than slow adaptation methods (Statement 14 and Statement 15 – 90% undecided, median ± IQR: 3 ± 0 for both).

**Fig. 1.**
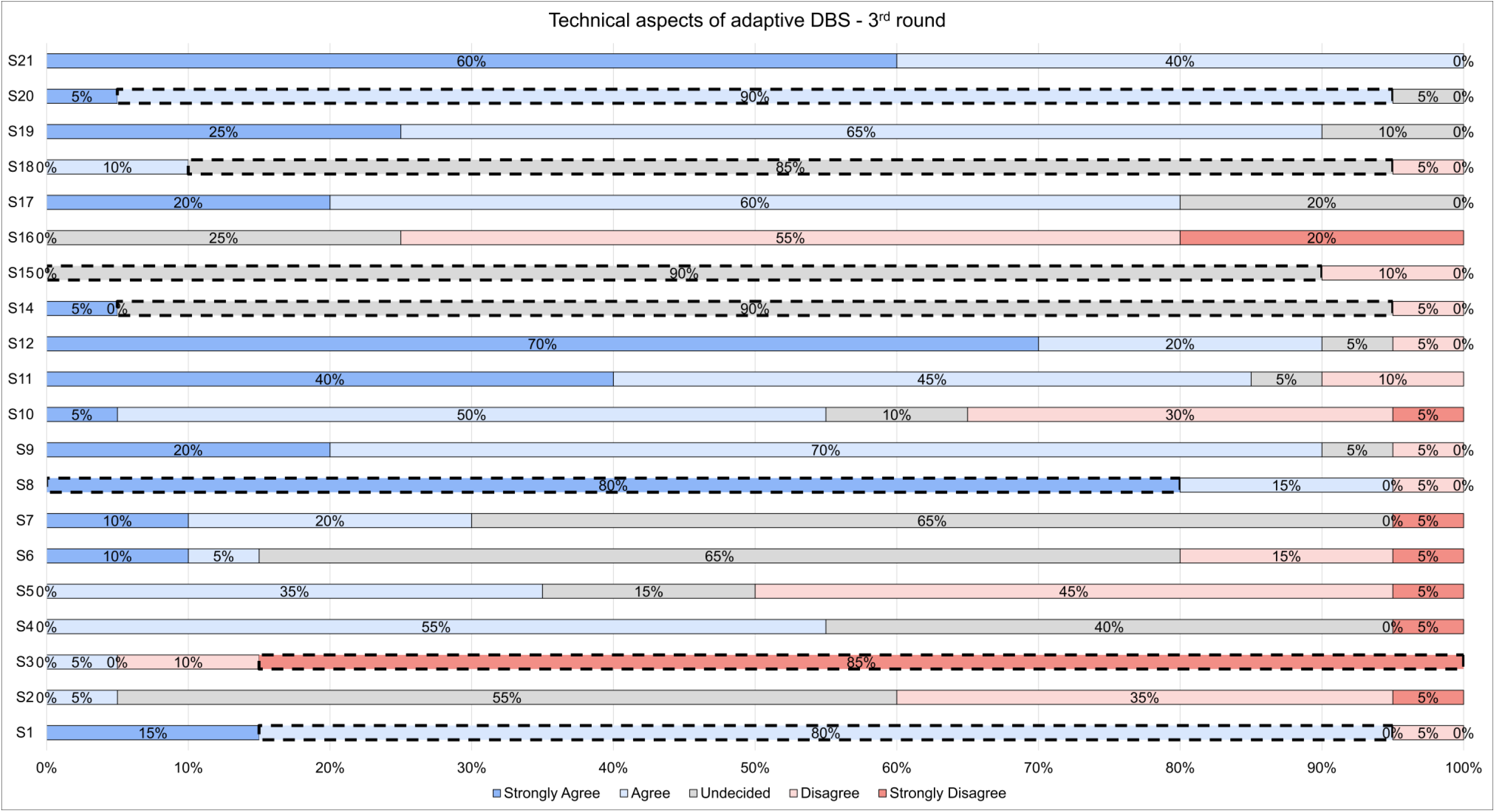
Percentage of agreement for the 21 statements on the technical aspects of adaptive DBS (Statement 1-21) among the Delphi Panel members, as result of the third round. A consensus was reached for Statement 1 (80% of the responses fell in the response label “Agree”), Statement 3 (85% of the responses fell in the response label “Strongly Disagree”), Statement 8 (80% of the responses fell in the response label “Strongly Agree”), Statement 14 (90% of the responses fell in the response label “Undecided”), Statement 15 (90% of the responses fell in the response label “Undecided”), Statement 18 (85% of the responses fell in the response label “Undecided”), and Statement 20 (90% of the responses fell in the response label “Agree”). DBS = Deep Brain Stimulation; S = statement.

**Fig. 2.**
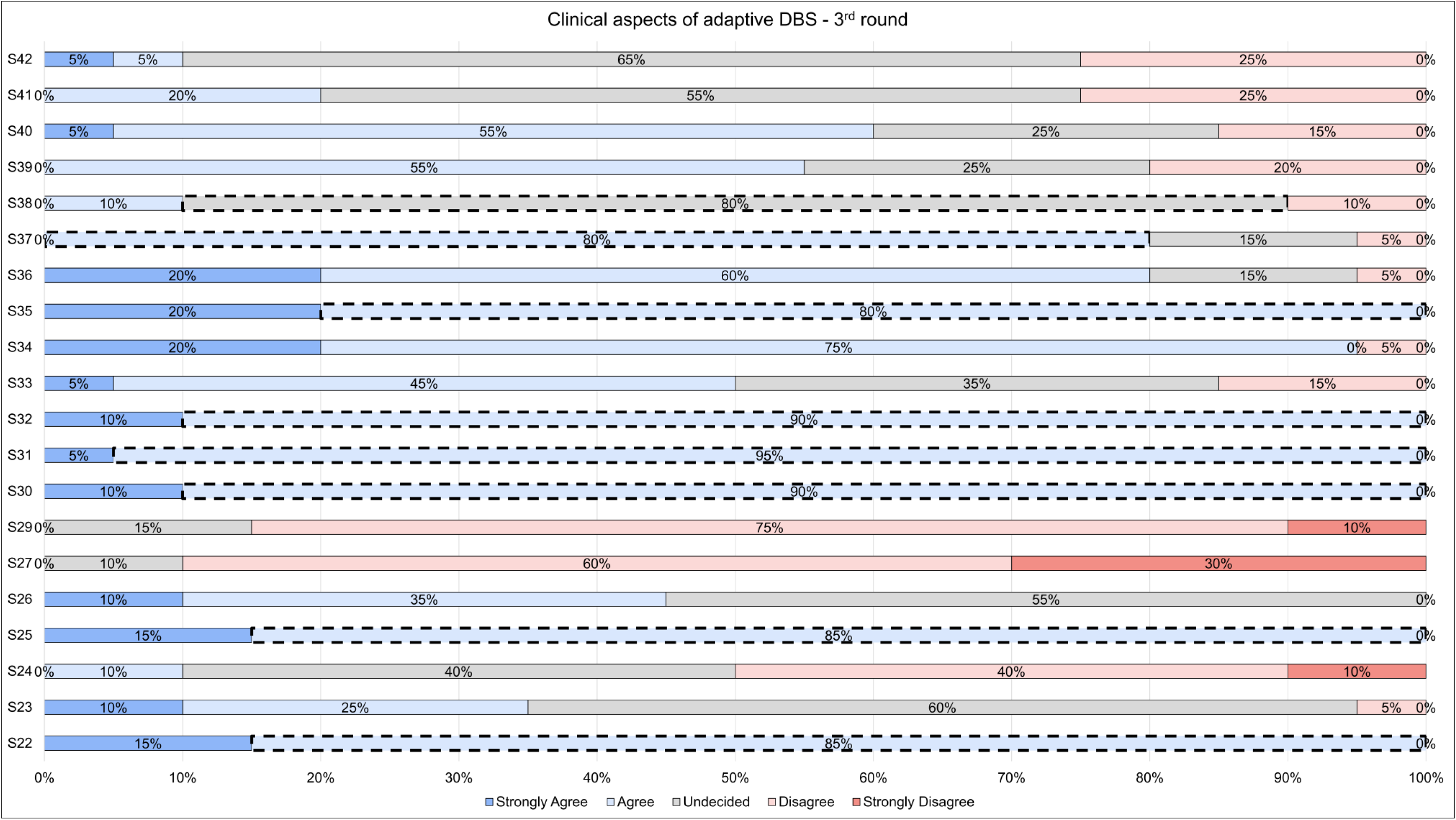
Percentage of agreement for the 21 statements on the clinical aspects of adaptive DBS (Statement 22-42) among the Delphi Panel members, as result of the third round. A consensus was reached for Statement 22 (85% of the responses fell in the response label “Agree”), Statement 25 (85% of the responses fell in the response label “Agree”), Statement 30 (90% of the responses fell in the response label “Agree”), Statement 31 (95% of the responses fell in the response label “Agree”), Statement 32 (90% of the responses fell in the response label “Agree”), Statement 35 (80% of the responses fell in the response label “Agree”), Statement 37 (80% of the responses fell in the response label “Agree”), and Statement 38 (80% of the responses fell in the response label “Undecided”). DBS = Deep Brain Stimulation; S = statement.

As for the 21 statements on the clinical aspects of aDBS, no consensus was reached after the first round (see eFigure 3 in Supplementary Materials). After the second, the panellists agreed on one statement (see eFigure 4 in Supplementary Materials), and other eight after the third round, for a total of 9 out of 21 statements (see fig.2). In particular, in the second round the panellists agreed on the use of aDBS technology also for tremor-dominant PD patients (Statement 28 – 80% agreed, median ± IQR: 4 ± 0). After the third round, an agreement was reached on the safety of aDBS technology (Statement 25 – 85% agreed, median ± IQR: 4 ± 0) and that it will enter clinical routine in 10 years (Statement 22 – 85% agreed, median ± IQR: 4 ± 0), with positive long-term impact for patients (Statement 35 – 80% agreed, median ± IQR: 4 ± 0), also for those with significant motor fluctuations before surgery (Statement 30 – 90% agreed, median ± IQR: 4 ± 0) and on cDBS treatment (Statement 31 – 95% agreed, median ± IQR: 4 ± 0), and for patients with significant dyskinesias on cDBS treatment (Statement 32 – 90% agreed, median ± IQR: 4 ± 0). Lastly, panellists agreed that aDBS might lead to a faster stable treatment response after the definition of stimulation settings (Statement 37 – 80% agreed, median ± IQR: 4 ± 0), but were uncertain if fast adaptation technology could lead to long term plastic changes (Statement 38 – 80% undecided, median ± IQR: 3 ± 0).

## 4. DISCUSSION

In this Delphi consensus study, 21 internationally recognized clinical and scientific experts in the DBS were asked to discuss current technical and clinical challenges related to aDBS development. Interestingly, out of the 42 open questions on aDBS proposed, a consensus was reached for 17, thus underlining the complexity and heterogeneity of the scenario and experiences as well as the general need of further research: experts agreed on a time frame of 10 years for aDBS to reach clinical practice whereas the time frame of 5 years did not achieve the agreement. To inform and support present adoption, the experience and knowledge gained so far suffice to reach a consensus regarding the safety of the adaptive approach and the potential benefits of aDBS. Experts in fact agreed that aDBS may lead to faster and more stable than cDBS treatment responses in selected patient populations, including tremor-dominant PD patients and those with motor fluctuations and dyskinesia on cDBS. Another important point related to the present scenario is the general agreement on the need of high level of expertise to manage aDBS, thus underlining a potential barrier to general adoption, but they also agreed that automatic programming can be safe if properly developed. The expert community remains uncertain regarding specific algorithms and their mechanisms of action, thus suggesting that future research and trials need to be directed towards the collection of data relevant both for understanding the neurophysiology of the adaptive approach and for identifying better biomarkers and the related stimulation patterns. Similarly, the possible combined benefits of aDBS and segmented electrodes remain unclear while there is general agreement on the fact that aDBS would not help in patients with electrodes that are not well positioned. Despite the high level of expertise, the lack of clinical and research evidence might have impaired the experts coming to a consensus on all the other aspects covered by the questions, both from the technical and the clinical point of view.

### 4.1. Technical aspects of aDBS

The panellists believe that despite the technological limitations of aDBS methodology, current pacemaker technology might be suitable to implement aDBS algorithms. Indeed, the recent development of pulse generators which are also able to record LFPs offers more options for optimising DBS therapy and aDBS algorithms.^37^ One of the main limitations of aDBS application in routine clinical care remains the uncertainty about which and how many signals could entirely represent patients’ clinical state and whether many of them need to be used together in multimodal algorithms.^8^ Most biomarkers have been identified with patients in “off stimulation”,^38^ but in the aDBS concepts, signals should be recorded in “on stimulation”. Therefore, the availability of devices able to record during stimulation is crucial to shed light on how to select the optimal personalised biomarker. While the most used closed-loop design (i.e., STN-LFP beta band as control signal to adjust for DBS amplitude) has been questioned,^15^ there is growing consensus that beta band is a fairly reliable biomarker.^39^ Several alternative approaches have been proposed (e.g., using cortical-subcortical gamma rhythm^40^), but no conclusive findings have been obtained yet.

The panellist acknowledged that a high level of expertise would be required to use aDBS. Indeed, currently, the programming phase of aDBS devices might require familiarity and higher technical skills (when compared with cDBS devices^41^), however the future algorithms will likely need to become more automated. This may suggest to industries to develop in the future simplified workflows or to provide adequate education to clinicians using aDBS. Still, clinicians will maintain a crucial role in assessing LFP recordings and their relationship to patient’s symptoms. As in any other new therapy, clinicians applying aDBS should keep the patient monitored to verify the persistence of an adequate control of symptoms over time and to modify pharmacological treatment if necessary. Adjustment of medications will likely be required independently of the type of stimulation (aDBS^42–44^ or cDBS^45^); however, combined effects of adaptive stimulation with medication might in selected cases decrease the risk of treatment-induced side effects like dyskinesia.

From the point of view of the level of automation in the approach, the experts agreed that automatic programming would be safe if stimulation intensity were constrained by combined upper and lower limits. The answer is in line with the need to avoid unpleasant side effects and an inadequate treatment of patients’ symptoms, especially for upper limits. However, many algorithms tested in clinical studies to date allow reduction of stimulation amplitude to zero when beta amplitude falls below a threshold, however, this could be modified in future fast aDBS algorithms.^39,42,44,46^

From a control algorithm point of view, the experts were uncertain about whether fast adaptation methods (movement-related) would be superior or inferior when compared to slow adaptation methods (drug-related). Indeed, beta activity can immediately trigger a brief increase in stimulation to shorten prolonged beta bursts^39,47^ or it can be smoothed over many seconds to serve as a medication state biomarker and then be used as feedback to drive stimulation.^44^ The way fast and slow adaptation algorithms have been implemented and studied, both reduced the total electrical energy delivered (TEED) over time by 50%, but while the first seems to reduce adverse effects on speech^48^ and to achieve a better control of bradykinesia and rigidity^44^ the latter seems to be more effective in reducing dyskinesias.^43^ These effects should be interpreted with great caution because of the paucity of cases and lack of independent validation. Indeed, speech was not systematically assessed for the “slow adaptation”, neither dyskinesias for the “fast adapting” algorithms. However, fast beta aDBS did also show the ability to adjust how often aDBS was triggered according to (slower) medication state, with stimulation becoming less frequent in the medication ON state. This suggests that “fast” aDBS algorithms can operate on both fast and slow timescales, and therefore could theoretically help medication induced dyskinesias.^49^ Currently, the lack of data does not allow to conclude differential benefits of both algorithms on side effects. Also, aDBS can possibly allow more TEED to be delivered, but with improved clinical efficacy and without inducing side effects;^40^ therefore, reduced TEED seems to be less of a critical outcome for DBS implementation, particularly with the advent of rechargeable devices.^50^

Panellists reached a consensus that the feasibility of aDBS for patients with suboptimally positioned electrodes was a limitation, meaning that it will likely not be effective. This expert opinion was in line with the evidence that the peak in beta activity is a feature of the motor part of the STN.^51^ Therefore, suboptimally positioned electrodes will not likely detect the LFPs needed to “adapt” aDBS to patients’ symptoms.

Similarly, the panellists were doubtful about the role of aDBS in facilitating the use of segmented electrodes, which may be used to widen the therapeutic window between efficacy and adverse effects by steering the field of stimulation.^52^ The experts did concede that segmented electrodes share with aDBS the common aim to “personalise” and shape stimulation electrical fields to single patients. Indeed, this technology increases spatial specificity while aDBS improves temporal specificity through the delivery of a dynamic stimulation that changes over time according to disease-related feedback.^52^ Theoretically, these two approaches could be complementary.

### 4.2. Clinical aspects of aDBS

The panellists shared an optimistic opinion in terms of development and applications of aDBS in clinical routine, and its potential ability to allow a faster and more stable treatment response in select patients. Indeed, despite the initial scepticism of parts of the medical community, the knowledge and technology in the field of aDBS have been growing.^53^ Also, recent technological advancements (e.g., directional leads^54^ or multiple stimulation methods^17,55^) may limit side effects and may serve to optimise for an individual symptom or symptoms.^8^

Another important point related to aDBS adoption is its safety, on which the panellists agreed. In addition to the surgical risks that to date are comparable to those of cDBS,^56^ concerns have been expressed in literature about the potential side effects of aDBS stimulation.^57^ Although no significant side effects have been reported so far,^58^ rapid changes of voltage or frequency induced by neurosignals could be unpleasant or even intolerable to patients in chronic stimulation. Thus, stimulation methods that balance ramp rates to avoid side effects and keep the stimulation therapeutic by responding in time to neurosignals changes are under study.^59^

One of the major potential advantages of aDBS is its ability to provide personalized therapy. The panellists agreed that aDBS is suitable both for PD patients experiencing motor fluctuations and dyskinesias before surgery or on cDBS, and for tremor-dominant PD patients. This consensus boosts the need of gaining more insights on the “precision medicine” potential of aDBS, i.e., investigating which patients are likely responders to stimulation, or which technology (e.g., which biomarker) is right for a specific patient.^60^ Beta frequency correlates more with rigidity/bradykinesia than resting tremor,^61,62^ while gamma activity, particularly finely-tuned gamma, has been associated with ON medication states and dyskinesia.^63,64^ Beta-driven aDBS might be less activated during levodopa-ON medication state (following beta suppression^38^) and hence reduces the likelihood of inducing levodopa-induced dyskinesia. Indeed, studies on aDBS in patients with PD and dyskinesia report good efficacy in reducing such symptom while guaranteeing a similar or even better control of cardinal symptoms of PD.^42,44,46^

Tremor can be detected from brain signals, either by the presence of lower frequency oscillations (3–7 Hz) or more accurately by combining multiple features from the whole-spectrum LFP.^65,66^ Additionally, several computational models have been recently developed to test the feasibility and efficacy of aDBS methods that modulate stimulation to control different biomarkers.^67,68^ In these cases, the best control may be provided by selecting between multiple controllers depending on context or patient symptoms (i.e., tremor or beta oscillations). Recent studies suggest a similar efficacy of aDBS both for tremor and bradykinesia dominant patients.^69,70^ Additionally, peripheral sensors may also be used for adaptive DBS for tremor.^71,72^

Major uncertainties remain on the mechanisms of action of aDBS: the experts were uncertain that fast adaptation technology could lead to long-term plastic changes. Although one might expect an effect close to what has been supposed for cDBS,^73^ whether aDBS might induce neuroplastic changes remains an open question due to the lack of evidence to support any opinion. Similarly, it is still to be determined what impact aDBS will have on the habituation phenomenon (i.e., the progressive loss of DBS benefit in time due to a decreased biological response of the neuronal networks^74^) that may in select cases threaten the effectiveness of cDBS in chronic conditions.^74^ However, some experts believe that habituation of DBS in the setting of PD is rare and that most of the worsening of symptoms is driven by PD progression.

### 4.3. Limitations

The consensus reached among experts as for the Delphi methods provides only the lowest level of evidence for making causal inferences.^75^ Therefore, the outcome of the present panel review cannot replace clinical judgments or original research, nor is it intended to define a standard of practice. Similarly, the feasibility of the consensus reached should be further debated and scientifically demonstrated – even more when considering stimulation targets commonly used for DBS (e.g., globus pallidus internus) not explored for aDBS. Rather, since our results aggregate the opinion of experts who could count on both personal expertise and scientific knowledge, they appear to be relevant in terms of current state of knowledge and future directions for research, even more for a field which is still at its infancy.

### 4.4. Conclusions

Despite experts agreed only partially on some technical aspects of aDBS, the panel concluded that aDBS will be routine in the mid-term. As for now, safety is a key aspect that reached agreement as well as the potential of aDBS to provide faster and more stable treatment response than cDBS, and in tremor-dominant PD patients and in those with motor fluctuations and dyskinesias. The expert panel also agreed that the neurophysiological mechanisms of aDBS, the best control strategy, and the relationship between this technology and other DBS-related innovations, such as segmented leads, are still to be investigated, thus orienting future research. Also, the current need of high level of expertise for the programming and management of aDBS patients represent a challenge that requires the coordination between research and industry, with automatic programming being an important development. In conclusion, the results of this Delphi consensus represent a step forward for aDBS to reach clinical adoption.

## Data Availability

All data produced in the present study are available upon reasonable request to the authors

## Contributors

M.G., A.F., J.K.K., A.A.K., A.L., M.O., A.P., L.T., and J.V. contributed to Conceptualisation and Methodology. J.O., G.D., P.S., S.L., R.M.F., G.T., C.G S., M.A.P., Y.T., A.S., P.L., C.H., and V.V.V. contributed to Methodology. M.G., S.M., N.V.M., S.O., T.B., and E.S. contributed to Data curation, Formal Analysis, Visualization and Writing – original draft. A.P. contributed to general study design, study ideation, discussion and supervision. All the authors contributed to Writing – review & editing, and accept responsibility for the decision to submit for publication.

