## Supplementary materials for "Adaptive Deep Brain Stimulation in Parkinson’s Disease: A Delphi Consensus Study"

**One eTable:**

1. Authors panellists in the Steering Committee (SC) and the Expert Panel (EP).

**Four eFigures**

1. Percentage of agreement for the 21 statements on the technical aspects of adaptive DBS (Statement 1-21) among the Delphi Panel members, as result of the first round.
2. Percentage of agreement for the 21 statements on the technical aspects of adaptive DBS (Statement 1-21) among the Delphi Panel members, as result of the second round.
3. Percentage of agreement for the 21 statements on the clinical aspects of adaptive DBS (Statement 22-42) among the Delphi Panel members, as result of the first round.
4. Percentage of agreement for the 21 statements on the clinical aspects of adaptive DBS (Statement 22-42) among the Delphi Panel members, as result of the second round.

**eTable 1. Authors panellists in the Steering Committee (SC) and the Expert Panel (EP).**

| Steering Committee (n=8) | Expert Panel (n=13) |
|---|---|
| Fasano Alfonso | Ostrem Jill L. |
| Krauss Joachim K. | Deuschl Günther |
| Kühn Andrea A. | Starr Philip A. |
| Lozano Andres M. | Little Simon |
| Okun Michael S. | Martinez-Fernandez Raul |
| Priori Alberto | Tinkhauser Gerd |
| Timmermann Lars | Gasca-Salas Carmen |
| Volkmann Jens | De Pedro Marta Del Álamo |
| · | Temel Yasin |
| · | Schnitzler Alfons |
| · | Limousin Patricia |
| · | Hamani Clement |
| · | Visser-Vandewalle Veerle |

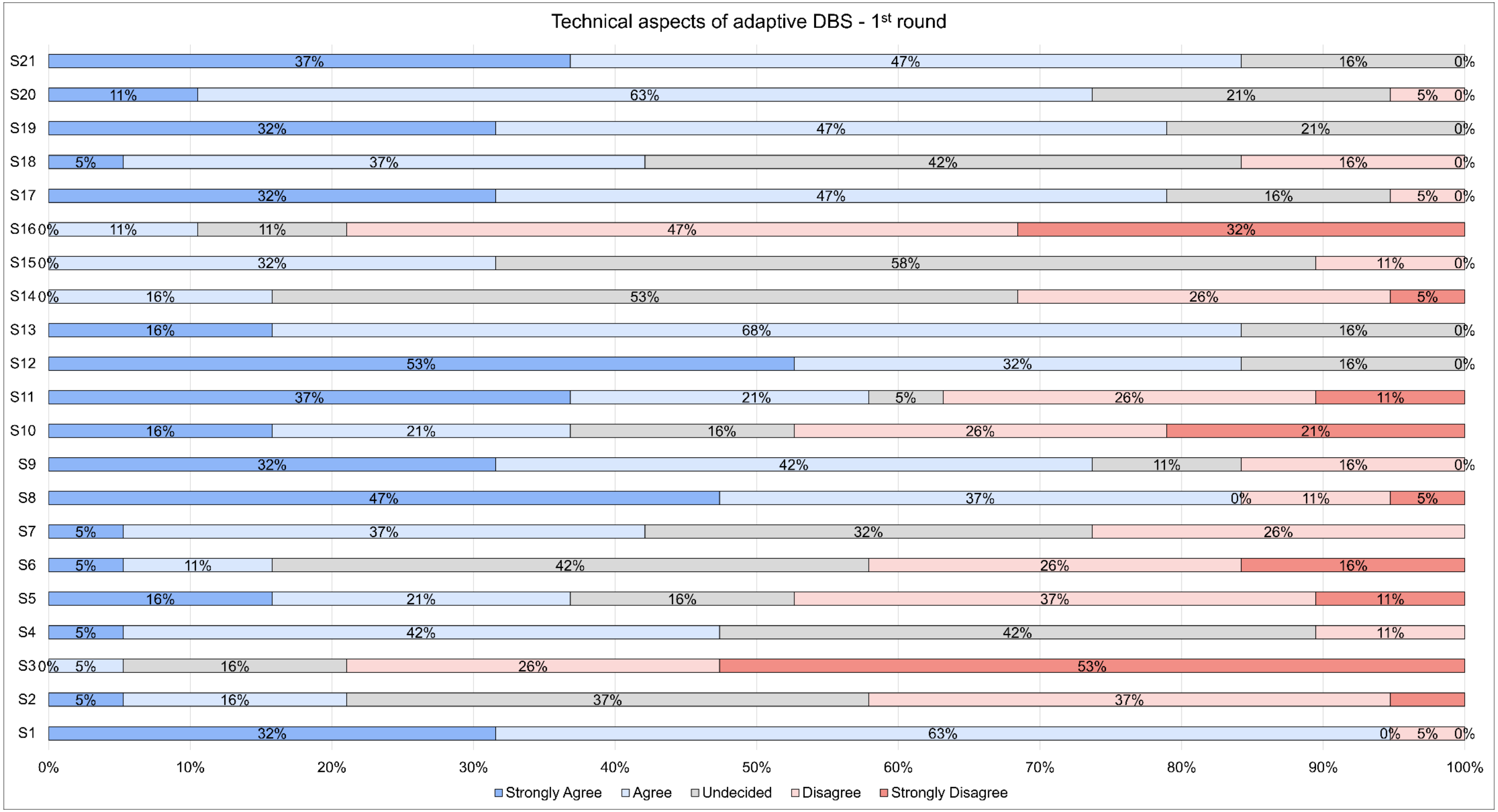


**eFigure 1. Percentage of agreement for the 21 statements on the technical aspects of adaptive DBS (Statement 1-21) among the Delphi Panel members, as result of the first round.** No statement reached a consensus (i.e. >80% of the responses fell in the same response label). DBS = deep brain stimulation; S = statement.

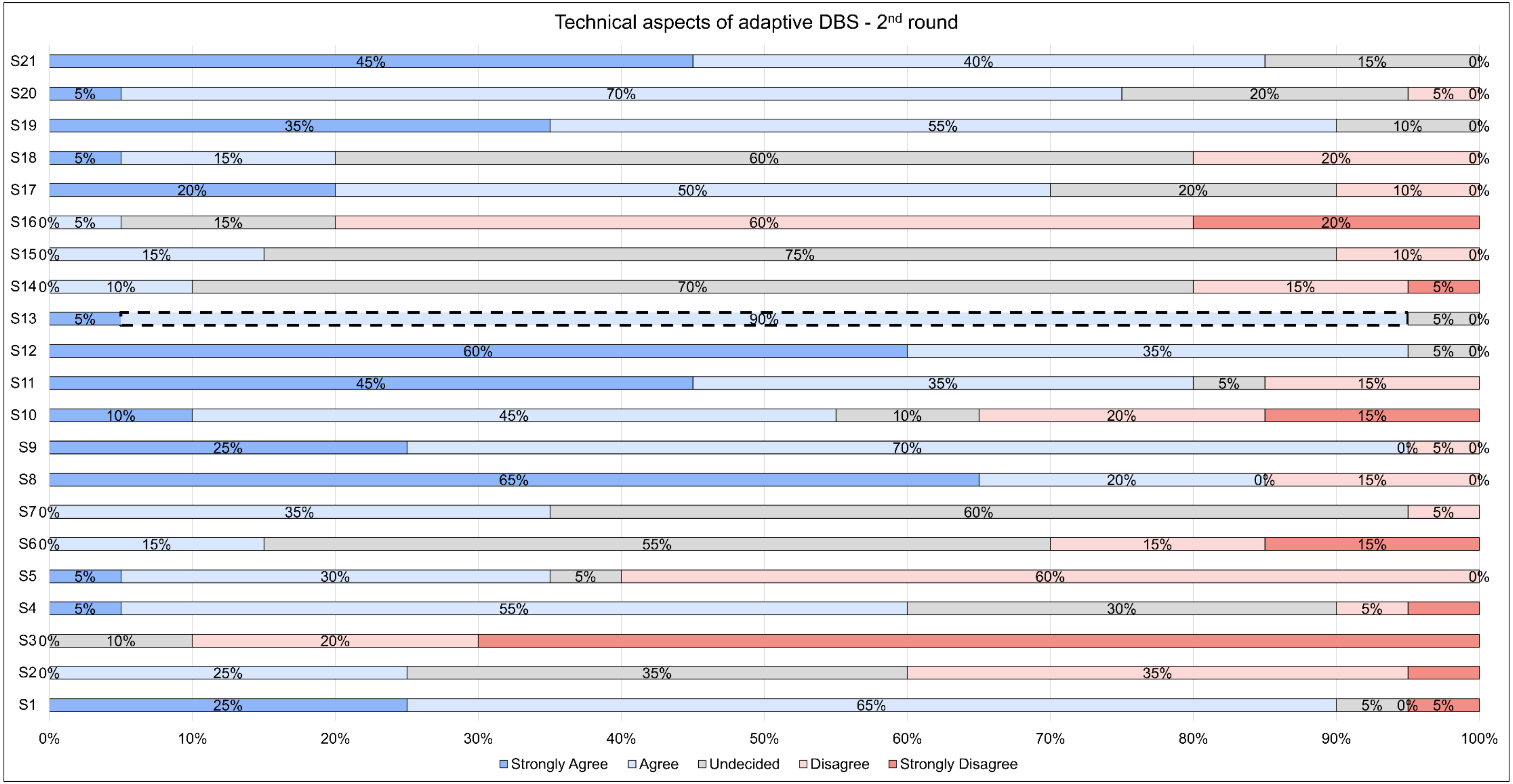


**eFigure 2. Percentage of agreement for the 21 statements on the technical aspects of adaptive DBS (Statement 1-21) among the Delphi Panel members, as result of the second round.** A consensus was reached for Statement 13 (90% of the responses fell in the response label "Agree"). DBS = deep brain stimulation; S = statement.

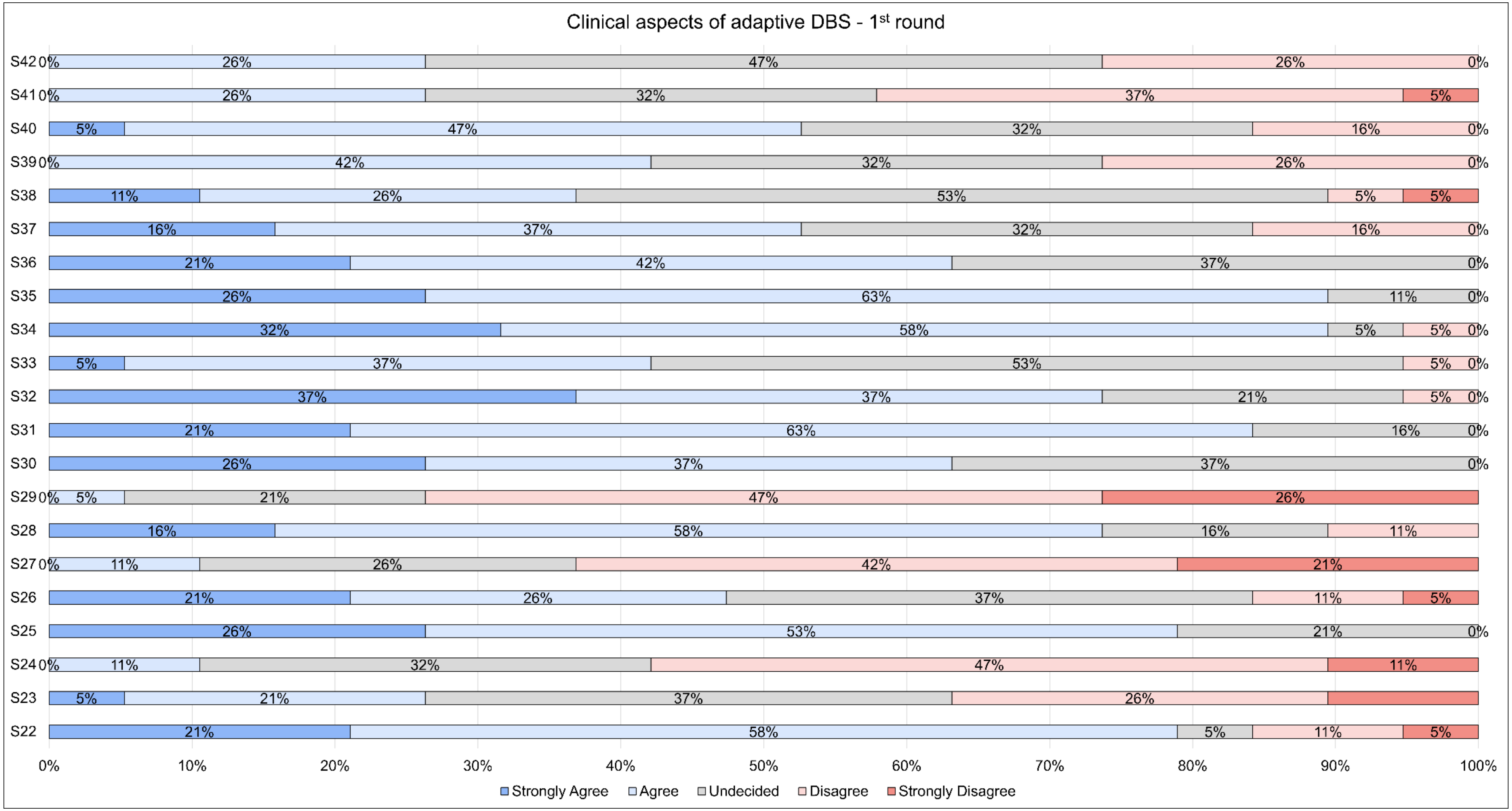


**eFigure 3. Percentage of agreement for the 21 statements on the clinical aspects of adaptive DBS (Statement 22-42) among the Delphi Panel members, as result of the first round.** No statement reached a consensus (i.e. >80% of the responses fell in the same response label). DBS = deep brain stimulation; S = statement.

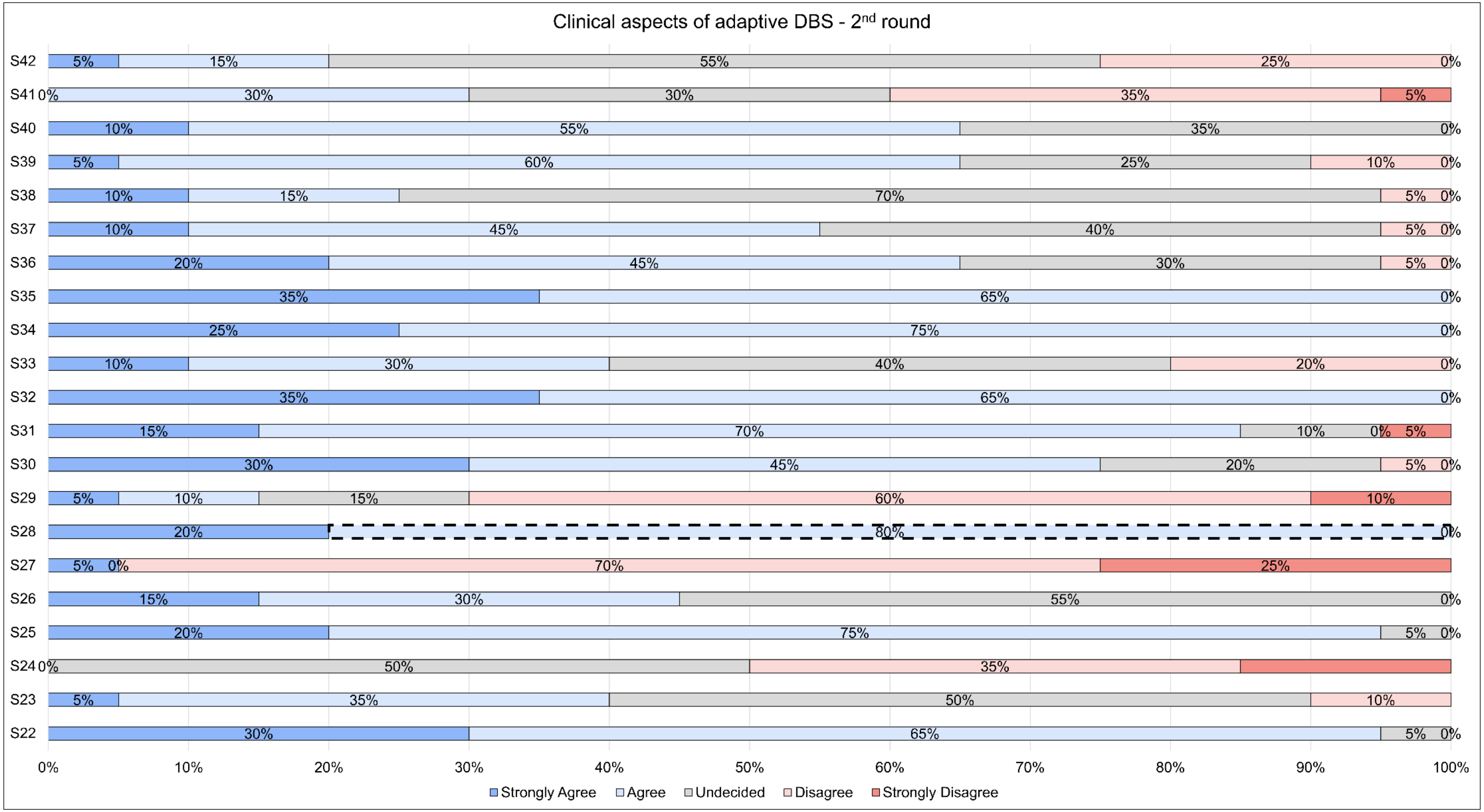


**eFigure 4. Percentage of agreement for the 21 statements on the clinical aspects of adaptive DBS (Statement 22-42) among the Delphi Panel members, as result of the second round.** A consensus was reached for Statement 28 (80% of the responses fell in the response label “Agree”). DBS = deep brain stimulation; S = statement.
